# Evolution of attitudes toward people with disabilities in healthcare practitioners and other occupations from 2006 to 2024

**DOI:** 10.1101/2025.03.11.25323797

**Authors:** Matthieu P. Boisgontier

**Affiliations:** Faculty of Health Sciences, University of Ottawa, Canada; Institut du Savoir Montfort, Hôpital Montfort, Ottawa, Ontario, Canada; Bruyère Health Research Institute, Ottawa, Canada; Perley Health Centre of Excellence in Frailty-Informed Care, Ottawa, Canada

**Keywords:** Attitude of Health Personnel, Bias, Disabled Persons, Healthcare Disparities, Prejudice, Professional-Patient Relations

## Abstract

**Purpose:** Healthcare practitioners have shown implicit and explicit attitudes that disfavor people with disabilities. This study aimed to describe how these attitudes evolved between 2006 and 2024 across clinicians, rehabilitation assistants, and other occupations.

**Methods:** In this comparative repeated cross-sectional study, data from 660,430 participants from Project Implicit were analyzed. Implicit attitudes were assessed using D-scores derived from the Disability Implicit Association Test. Explicit attitudes were assessed using a Likert scale. Generalized additive models were conducted to test the evolution of attitudes over time.

**Results:** Explicit attitudes toward people with disabilities became less unfavorable over time, following a linear pattern. No such effect was found for implicit attitudes. However, non-linear interactions between time, occupation group, and sex suggest a complex effect of time on attitudes that should be interpreted in the context of each specific combination of occupation group and sex, rather than assuming a uniform trend. Attitudes were less favorable toward people with physical disabilities than general disabilities.

**Conclusions:** The contrast between evolution of implicit and explicit attitudes suggests that implicit bias remains resistant to change despite improving explicit consideration of people with disabilities. Knowledge of these patterns may inform training programs to reduce bias in healthcare and beyond.

## INTRODUCTION

Human behavior is influenced by a tendency to evaluate entities from the environment with some degree of favor or disfavor.^1,2^ This tendency results in attitudes toward various social groups and behaviors, such as race,^3^ age,^4^ and physical activity.^5^ These attitudes can be explicit and consciously controlled,^6^ or implicit (i.e., automatic),^7^ reflecting traces of past experience that remain introspectively unidentified.^8^

Understanding health practitioners’ attitudes toward people with disabilities is essential for reducing potential biases in care, especially given the historical framing of disability through a deficit lens.^9^ This framework that conceptualizes disability as an abnormality that needs to be normalized to conform to societal ideals of “normalcy”^9^ has been embedded in healthcare practices for decades.^10^ Although this deficit framework has been criticized for overlooking the importance of inclusion and accessibility,^9,10^ studies suggest it continues to affect healthcare professionals’ attitudes through broader systemic and cultural influences.^10^ This deficit framework aligns with the biomedical model of disability, which contrasts with the social model. The latter conceptualizes disability as the result of structural, institutional, and attitudinal barriers rather than individual impairment.^11^ While the social model has been instrumental in shifting focus toward environmental and social contributors to disability, it has been criticized for minimizing the significance of physical and psychological factors.^12^ In response, more integrated models have emerged, positing that disability arises from the interaction between bodily differences and social context.^9,11,12^ One such model is the World Health Organization’s (WHO) International Classification of Functioning, Disability and Health (ICF),^13^ which offers a biopsychosocial framework. Within these integrative models, attitudes toward disability may act as a key mechanism through which environmental factors, such as cultural norms and institutional structures, influence access to care, clinical decision-making, and health-related outcomes.

Recent large-scale studies have shown that healthcare professionals exhibited less favorable implicit and explicit attitudes toward people with disabilities than toward people without disabilities (Fig. 1A-C).^14–16^ Specifically, a study of healthcare professionals (including clinicians, physiotherapy assistants, occupational therapy assistants, nursing assistants, home health assistants, technologists, technicians, and other healthcare support personnel) showed a slight explicit preference for people without disabilities and a moderate implicit preference.^14^ Another study focusing on rehabilitation assistants showed similar results.^15^ Further, a comparative study focusing on attitudes toward *physical* disability (Fig. 1C-D) showed that the implicit attitudes of clinicians and rehabilitation assistants were similar to those in other occupations.^16^ However, no studies have tested whether these attitudes have evolved in recent decades across occupational groups, particularly in healthcare. Addressing this knowledge gap is essential to understand whether and how attitudes are shifting. This knowledge would inform efforts to promote inclusion and equity in healthcare and beyond.

**Figure 1.**
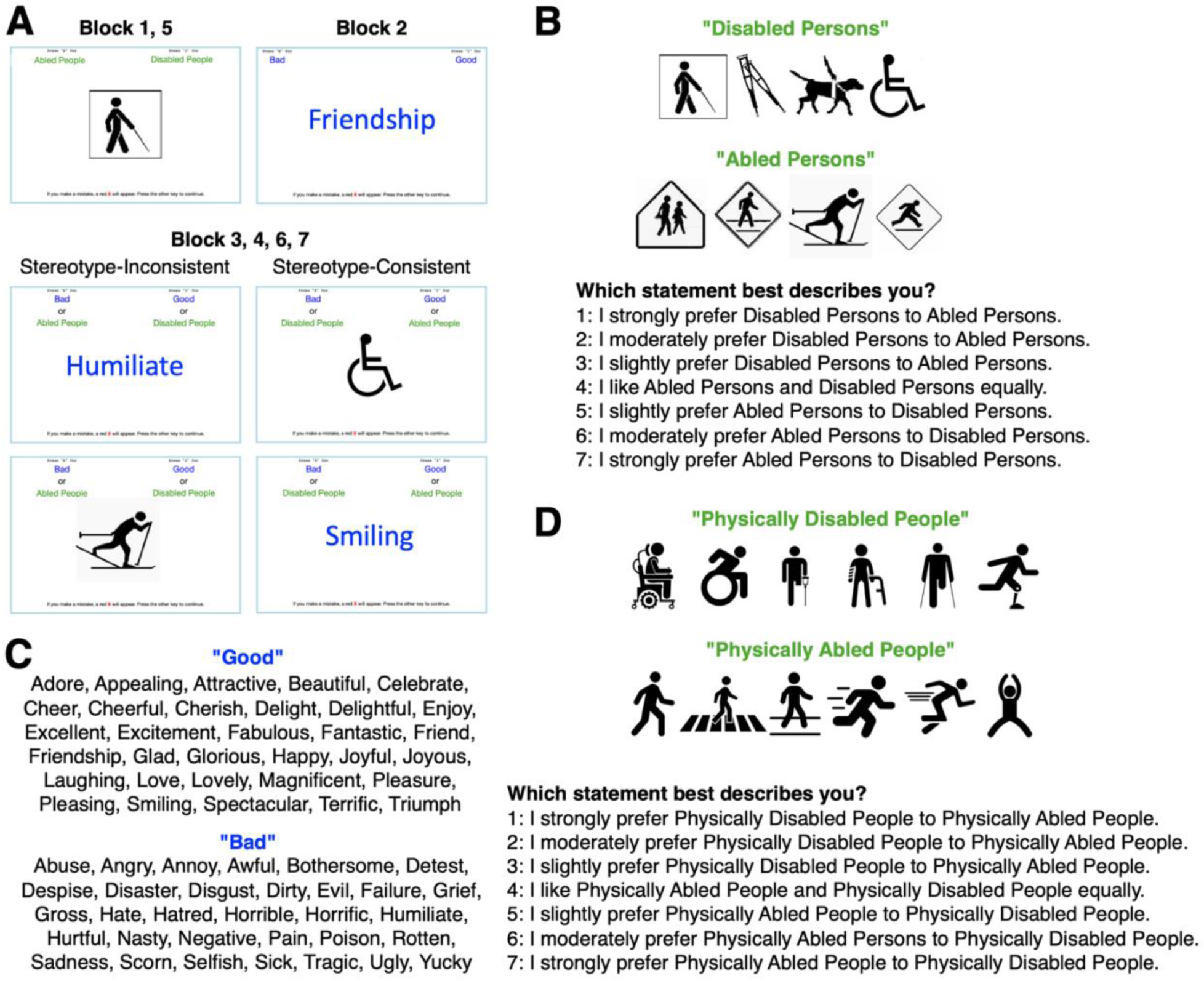
Assessment of implicit and explicit attitudes. (A) Screenshots of the Disability Implicit Association Test. (B) Images and questions used to assess attitudes toward people with *general* disabilities. (C) Words used for the evaluative attributes (“good” and “bad”). (D) Images and questions used to assess attitudes toward people with *physical* disabilities.

The objective of this study was to describe how implicit and explicit attitudes toward people with disabilities evolved between 2006 and 2024 among clinicians, rehabilitation assistants, and other occupation groups. Since “understanding scientific findings in the context of sex—be they similarities, differences, and/or complex nuances—is crucial for correctly applying research-derived knowledge”,^17^ sex was included as a potential moderator of this relationship.

## METHODS

### Participants

This study is based on two Implicit Association Test (IAT) datasets collected through the publicly accessible Project Implicit website (https://implicit.harvard.edu/implicit/selectatest.html) and made available on the Open Science Framework (OSF) under the CC0 1.0 Universal License.^18^ The website allows any adult aged 18 years of age or older to voluntarily participate and measure their implicit and explicit attitudes toward people with disabilities, and to answer demographic questions (e.g., age, sex). Participants were informed that data without directly identifying information would be made publicly available for research purposes.

### Disability Implicit Association Test

#### Assessment of Implicit Attitudes

The Disability IAT assesses implicit attitudes toward people with and without disabilities. In other words, this test measures the strength of automatic associations between the target concepts (i.e., people with vs. without physical disabilities) and evaluative attributes (i.e., good vs. bad). The underlying principle is that participants respond more quickly when strongly associated categories share the same response key, reflecting implicit associations. The Disability IAT shows robust internal consistency (Cronbach’s alpha = 0.85),^19^ moderate test-retest reliability (r = 0.50),^20^ small predictive validity (r = 0.09),^19^ and moderate incremental predictive validity, significantly predicting 10 out of 17 outcomes in structural equation models beyond what was accounted for by explicit attitudes.^19^

#### General (2006-2021) and Physical Disability (2022-2024)

The type of disability assessed through the Disability IAT on the Project Implicit website has changed over time. From 2006 to 2021, the IAT measured attitudes toward *general* disability. Since 2022, the IAT measures attitudes toward *physical* disability. The two versions differ in two ways. First, the 2006-2021 version included four images representing disability: a guide dog, a person with a cane, crutches, and the International Symbol of Access (wheelchair) (Fig. 1A-B). In contrast, the 2022–2024 version included six images, all of which represent physical disability (Fig. 1D). Second, the 2006-2021 version used a 7-point Likert scale to assess explicit attitudes toward “abled persons” and “disabled persons” (Fig. 1B), whereas the 2022–2024 version assessed explicit attitudes toward “physically disabled people” and “physically abled people” (Fig. 1D). The set of 16 words for each attribute was the same across versions (Fig. 1C). Detailed procedures are described in Suppl. Material 1.

### Outcome Variables

#### Implicit Attitudes

Implicit attitudes toward people with and without disabilities were assessed using the D-score measure (Suppl. Material 1),^21^ which is based on participants’ performance on blocks 3, 4, 6, and 7 of the Disability IAT (Fig. 1A). This measure divides the difference between the mean response latency on the stereotype-consistent trials (e.g., “disabled people” paired with “bad” and “abled people” paired with “good”) and the mean response latency on the stereotype-inconsistent trials (e.g., “disabled people” paired with “good” and “abled people” paired with “bad”) by the standard deviation of all the latencies across the four blocks:

Error trials were included. Trials with response latencies below 400 ms and above 10,000 ms were excluded to reduce the influence of random or disengaged responses, and participants with more than 10% of trials below 300 ms were excluded to ensure data validity.^16^

#### Explicit Attitudes

Explicit attitudes were assessed using a 7-point Likert scale in which participants rated their preference for people with or without disabilities. A score of 1 indicated a strong preference for people with disabilities, 4 indicated no preference, and 7 indicated a strong preference for people without disabilities (Suppl. Material 2).

### Explanatory Variables

*Occupation.* Participants’ occupation was determined by their response to the item: “Please select the most appropriate occupation category”. Participants who selected “Healthcare – Diagnosing and treating practitioners” (e.g., medical doctor, physiotherapist, nurse, dentist) were categorized as clinicians, while those who selected “Healthcare – Occupational and physical therapist assistants” were categorized as rehabilitation assistants. All other occupations were categorized as “other occupations”. A complete list of occupation categories is available in Suppl. Material 3.

*Time.* A continuous variable was derived by combining the year, month, and day of data collection into a single date format (YYYY-MM-DD). This date was then converted into a numeric variable representing the number of days since 1970-01-01 to facilitate its use as a continuous variable.

*Age.* Age was treated as a continuous variable determined by the question asking participants their age in years. If this question was not asked, age was calculated as the difference between the year of data collection and the participant’s year of birth. As the focus of our study was on occupation, participants under the age of 20 and over the age of 70 were excluded.

*Sex.* Participants’ sex was determined by the question “What sex were you assigned at birth, on your original birth certificate?” If this question was not asked, we used the answer to a gender-related question that included male and female as response options.

*Disability Type.* A categorical variable was created to account for the two versions of the Disability IAT (general vs. physical).

### Statistical Analyses

All analyses were conducted in R (version 4.4.1).^22^ To examine the evolution of implicit and explicit attitudes toward people with disabilities across different occupational groups and sexes, between 2006 and 2024, we fitted two separate generalized additive models using the mgcv package (version 1.9-1). These models robustly capture potential non-linear relationships between outcomes and explanatory variables,^23,24^ as evidenced in studies investigating the effects of age on brain structure and function.^25,26^

The outcome of the first model was the D-score representing implicit attitudes toward people with disabilities. The explanatory variables included time, occupation group (clinicians, rehabilitation assistants, other occupations), sex (male, female), age, explicit attitudes (Likert score), and disability type (general, physical). The interaction between time, occupation group, and sex was modeled using the tensor product smooth function. In this interaction, time was modeled with a cubic regression spline to capture potential non-linear relationships between time and implicit attitudes toward people with disabilities. This spline provides flexibility to account for complex temporal trends, recognizing that the effect of time on attitudes may not follow a linear pattern. Occupation group and sex were treated as random effects, recognizing that different occupational groups and sexes may have distinct baseline levels of attitudes toward disability without requiring a smooth or continuous transition between their categories. The second model estimated explicit attitudes (Likert score) using the same structure, with D-score replacing explicit attitudes as an explanatory variable. Continuous explanatory variables were standardized. The significance of the smooth terms was evaluated using F-tests, while Wald tests were used to assess the significance of the fixed effects. The significance level (α) was set at 0.05. The effective degrees of freedom (edf) from the generalized additive models were used to assess the complexity of the smooth functions, with higher complexity typically reflecting greater non-linearity: 1 was considered to indicate a linear relationship], 1–5] mild complexity], 5–10] moderate complexity, and >10 high complexity.

## RESULTS

### Descriptive Results

A total of 660,430 participants from three occupation groups were included: clinicians, rehabilitation assistants, and participants in other occupations. Implicit attitudes toward people with disabilities were broadly similar across occupation groups. Explicit attitudes were slightly more unfavorable among clinicians and slightly less unfavorable among rehabilitation assistants than in participants with other occupations. Clinicians and participants in other occupations were older than rehabilitation assistants. The proportion of female participants was highest among rehabilitation assistants. Most participants completed the version of the IAT assessing general disability, with physical disability assessed in a smaller subset across all groups. Descriptive statistics for all groups are summarized in Table 1 and a detailed breakdown of the number of participants per year and occupational group is presented in Supplementary Material 4.

**Table 1.** Descriptive characteristics of the study sample by occupation group.

| Exposures | Clinicians<br>(n = 20,405) | Rehabilitation Assistants<br>(n = 9357) | Other Occupations<br>(n = 630,668) |
| --- | --- | --- | --- |
|  | Mean ± SD | Mean ± SD | Mean ± SD |
| **Implicit Attitudes** (D-score) | 0.53 ± 0.44 | 0.51 ± 0.45 | 0.53 ± 0.45 |
| **Explicit Attitudes** (Likert score) | 4.44 ± 0.82 | 4.27 ± 0.76 | 4.34 ± 0.86 |
| **Age** (years) | 33.6 ± 11.9 | 28.6 ± 9.4 | 33.6 ± 12.3 |
|  | Count (%) | Count (%) | Count (%) |
| **Female Participant** | 14,056 (68.9) | 7863 (84.0) | 470,520 (67.9) |
| **Male Participant** | 6349 (31.1) | 1494 (16.0) | 160,148 (25.4) |
| **General Disability** | 15,235 (74.7) | 6552 (70.0) | 465,097 (73.8) |
| **Physical Disability** | 5170 (25.3) | 2805 (30.0) | 165,571 (26.2) |

### Implicit Attitudes

The generalized additive model examining implicit attitudes toward people with disabilities, as measured by the D-score, explained 4.9% of the variance (adjusted R² = 0.049).

Analysis of the fixed effects (Fig. 2A) showed no evidence of a linear effect of time on implicit attitudes (b = –1.9 × 10^−6^, 95% CI [-5.7 × 10^−6^ to 1.9 × 10^−6^]; *P* = .329). Older (b = 5.3 × 10^−3^, 95% CI [5.2 × 10^−3^ to 5.4 × 10^−3^]; *P* < 2 × 10^−16^) (Fig. 3A) and male participants (b = –0.116, 95% CI [-0.133 to –0.099]; *P* < 2 × 10^−16^) showed more unfavorable implicit attitudes toward people with disabilities than younger and female participants, respectively. Compared to participants in other occupations, rehabilitation assistants had more unfavorable implicit attitudes toward people with disabilities (b = 0.036, 95% CI [0.014 to 0.058]; *P* = 1.2 × 10^−3^), while no difference was observed for clinicians (b = 8.6 × 10^−4^, 95% CI [-0.019 to 0.020]; *P* = .931). Implicit attitudes were more unfavorable toward people with physical disabilities compared to general disabilities (b = 9.0 × 10^−3^, 95% CI [4.4 × 10^−3^ to 1.4 × 10^−2^]; *P* = 1.3 × 10^−4^) (Fig. 3A). Explicit attitudes were positively associated with implicit attitudes (b = 0.065, 95% CI [0.064 to 0.066]; *P* < 2 × 10^−16^), indicating that weaker explicit preferences for people with disabilities were associated with weaker implicit preferences (Fig. 3A).

**Figure 2.**
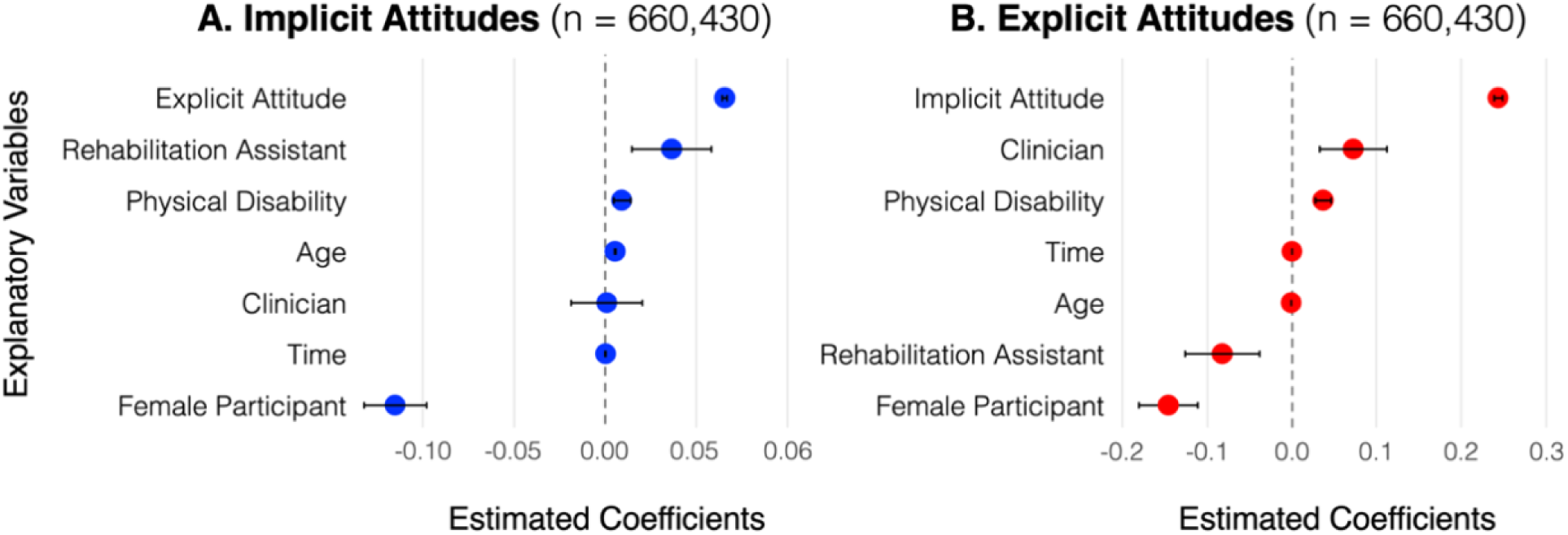
Estimated coefficients of the fixed effects from the generalized additive models examining the association of explanatory variables with implicit. (A) and explicit attitudes (B) toward people with disabilities. Positive coefficients indicate less favorable attitudes, whereas negative coefficients indicate more favorable attitudes. For the categorical variables, the reference categories are “other occupation”, “male participant”, and “general disability”. The figure shows the estimated coefficients (points) and 95% confidence intervals (error bars). For clarity, continuous variables (i.e., age, explicit attitudes, implicit attitudes, time) are presented in their original units (i.e., were not standardized).

**Figure 3.**
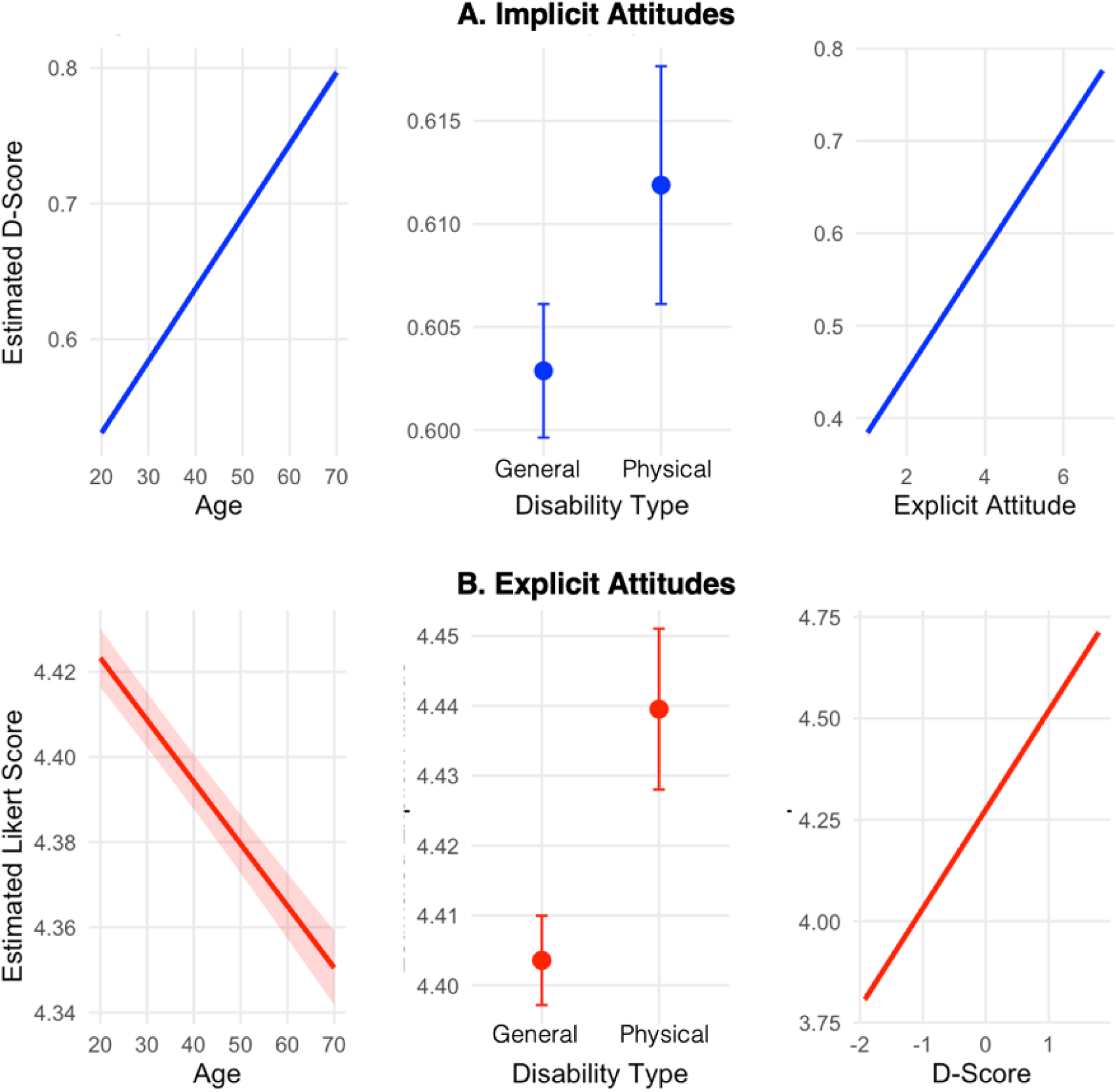
Implicit (A) and explicit attitudes (B) as a function of age (left), disability type (middle), and the other type of attitudes (right), as estimated by the generalized additive models. The left and right panels show smooth effects with 95% confidence intervals (shaded areas), while the middle panel displays estimated means with 95% confidence intervals (error bars). Predictions are adjusted for time, occupation group, and sex.

Beyond the fixed effects, results showed an interaction between time, occupation group, and sex, modeled using a tensor product smooth (F = 2.922; edf = 11.88; *P* < 2 × 10^−16^) (Fig. 4A).

**Figure 4.**
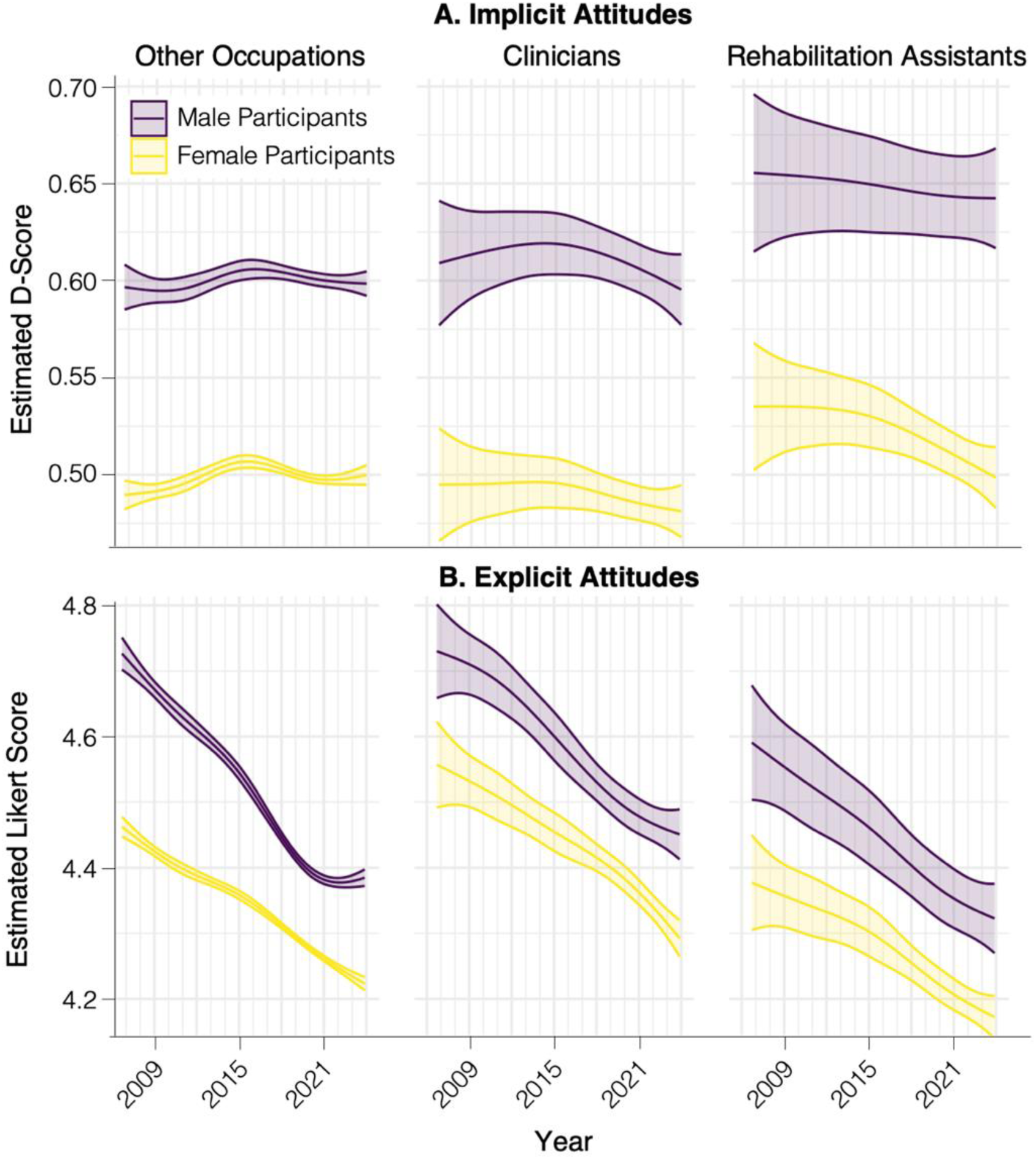
Results of the three-way interactions showing implicit. (A) and explicit attitudes (B) over time as a function of occupation group and sex, as estimated by the generalized additive models. Solid lines represent estimated attitudes, and shaded areas indicate 95% confidence intervals.

This result indicated that the relationship between time and implicit attitudes toward people with disabilities varied by occupation group and sex, following a highly complex, non-linear pattern. The statistical significance and complexity of this smooth term suggested that these combined effects may not be adequately described by simple linear relationships.

### Explicit Attitudes

The generalized additive model examining explicit attitudes toward people with disabilities, as measured by the Likert score, explained 2.9% of the variance (adjusted R² = 0.029). Analysis of the fixed effects (Fig. 2B) showed that explicit attitudes toward people with disabilities became less unfavorable over time (b = –4.2 × 10^−5^, 95% CI [-5.0 × 10^−5^ to –3.4 × 10^−5^]; *P* < 2 × 10^−16^). In Fig. 2B, the effect size of time appears small because the unit of measurement is a day. However, this seemingly small daily effect accumulates over a 19-year period, resulting in a more substantial cumulative effect. Older (b = –1.5 × 10^−3^, 95% CI [-1.6 × 10^−3^ to –1.3 × 10^−3^]; P < 2 × 10^−16^) (Fig. 3B) and female participants (b = –0.147, 95% CI [-0.181 to –0.112]; P < 2 × 10^-16^) expressed less unfavorable explicit attitudes toward people with disabilities compared to younger and male participants, respectively. Compared to participants in other occupations, clinicians reported more unfavorable explicit attitudes (b = 0.072, 95% CI [0.033 to 0.111]; P = 3.4 × 10^−4^), while rehabilitation assistants reported less unfavorable explicit attitudes (b = –0.083, 95% CI [-0.127 to –0.038]; P = 2.5 × 10^−4^). Explicit attitudes were more unfavorable toward people with physical disabilities than toward people with general disabilities (b = 0.036, 95% CI [0.027 to 0.045]; *P* = 1.2 × 10^−14^) (Fig. 3B). Explicit attitudes were positively associated with implicit attitudes (b = 0.243, 95% CI [0.239 to 0.248]; *P* < 2 × 10^−16^) (Fig. 3B).

Beyond the fixed effects, results showed and interaction between time, occupation group, and sex, modeled using a tensor product smooth (F = 13.95; edf = 14.56; *P* < 2 × 10^−16^) (Fig. 4B). This result indicated that the relationship between time and explicit attitudes toward people with disabilities varied by occupation group and sex, following a highly complex, non-linear pattern. The statistical significance and complexity of this smooth term suggested that these combined effects may not be adequately described by simple linear relationships.

## DISCUSSION

### Main Findings

The present study examined the evolution of implicit and explicit attitudes toward people with disabilities between 2006 and 2024 among clinicians, rehabilitation assistants, and individuals in other occupations, considering sex as a moderating factor, and using a large-scale dataset (n = 660,430). Results showed no evidence suggesting a linear effect of time on implicit attitudes, while explicit attitudes became less unfavorable toward people with disabilities over time. However, non-linear interactions between time, occupation, and sex suggested that the effect of time on attitudes was complex and should be interpreted within the context of each specific combination of occupation group and sex, rather than assuming a uniform trend.

Overall, male participants consistently showed more unfavorable attitudes toward people with disabilities than female participants. Implicit and explicit attitudes were more unfavorable toward people with physical disabilities than people with general disabilities. Older age was associated with more unfavorable implicit attitudes but less unfavorable explicit attitudes toward people with disabilities. Rehabilitation assistants exhibited more unfavorable implicit attitudes than other occupation groups, whereas clinicians did not significantly differ. In contrast, explicit attitudes were less unfavorable among rehabilitation assistants and more unfavorable among clinicians than among participants in other occupation groups.

### Comparison with the Literature

Consistent with prior research, our results showed a moderate implicit preference and a slight explicit preference for people without disabilities, replicating known patterns in the literature.^14–16^ They also reinforce evidence that male healthcare practitioners exhibit more unfavorable attitudes toward people with disabilities than female practitioners.^14,16^

Building on a previous study that focused on physical disability,^16^ we compared implicit and explicit attitudes across clinicians, rehabilitation assistants, and individuals in other occupations. Unlike the previous study, this study included data on general disability. Overall, results were consistent across both studies. Clinicians showed more unfavorable explicit attitudes toward people with disabilities than participants in other occupations. In contrast, rehabilitation assistants showed less unfavorable explicit attitudes. In terms of implicit attitudes, no statistical differences were found between clinicians and participants in other occupations in either study. However, our results diverge in showing that rehabilitation assistants exhibited more unfavorable implicit attitudes, a pattern not observed in the previous study focusing on physical disability. One possible explanation for this discrepancy is that rehabilitation assistants may interact more frequently with people with disabilities in ways that emphasize functional limitations, potentially reinforcing implicit biases. In contrast, clinicians, such as physiotherapists and physicians, may approach disability from a more long-term, progress-oriented perspective centered on recovery and broader health outcomes, which may buffer against the development of stronger implicit biases.

Result showed that older participants displayed more unfavorable implicit attitudes but less unfavorable explicit attitudes toward people with disabilities. Although our design precludes causal interpretations, we hypothesize that this divergence may reflect generational differences. Older adults may have been more exposed to the deficit framework, leading to the persistence of negative implicit associations. Over time, however, they may have consciously updated their beliefs to align with contemporary values of inclusion and accessibility, resulting in more favorable explicit attitudes. This divergence suggests that, while explicit beliefs may shift with social norms, implicit attitudes may be more resistant to change.

Results showing that both implicit and explicit attitudes were more unfavorable toward people with physical disabilities than toward people with general disabilities suggest that visibility and perceived severity may influence bias. Therefore, future research should analyze attitudes by disability type.

### The Need for a Multimethod Assessment of Attitudes

It is important to acknowledge that the implicit and explicit attitude measures used in this study rely on evaluative concepts, such as “preference” or “bad” vs. “good”, that may reflect deficit-based views of disability by positioning it as a characteristic to be judged. In addition, these measures do not fully capture the nuanced and complex nature of disability. However, improving our understanding of attitudes, even through imperfect measures, is essential to improving healthcare and society. To address these limitations, future studies should incorporate these tools within a broader, multimethod approach to assessing disability bias.^28^ This approach could include approach-avoidance tasks,^27^ explicit self-report questionnaires, behavioral observations, physiological and neural measures, as well as qualitative interviews.^28^

### Educational Strategies for Reducing Bias Toward People with Disabilities

Although explicit attitudes toward people with disabilities have become more favorable over the years, implicit biases remained relatively stable, highlighting the need for more effective educational strategies. Interventions have shown to be most effective when they combine information delivery with relational or experiential learning, especially through direct or indirect contact with disabled people.^29^ Such multi-component approaches improve empathy, comfort, and understanding more than information alone.^29^ Further, lasting change in implicit attitudes requires sustained exposure to counter-stereotypical exemplars and systemic inclusion of disability across curricula, pedagogy, and placements.^30^ Without such environmental redesign and experience repetition, interventions are unlikely to yield lasting change in attitudes toward people with disabilities.^30^ Therefore, educational systems not only need to teach about bias but also to create conditions in which inclusive interactions become the norm, ultimately transforming automatic evaluative processes and behaviors.”

### Limitations

Several limitations should be considered. First, the study relied on observational data from a repeated cross-sectional design, which precludes causal inference. Therefore, observed changes in attitudes over the years could not be attributed to specific causes. Future studies should include longitudinal designs and randomized controlled trials. Second, the R^2^ values in our models were statistically significant but small, indicating that the observed group differences and associations explained only a modest portion of the variance in implicit and explicit attitudes. These small but significant effect sizes align with previous studies on attitudes using large datasets and reflect the complexity of attitudes, which are influenced by a wide range of individual and contextual factors. Nonetheless, small effects can be meaningful in real-world contexts, particularly when they accumulate over the years to influence behaviors. Therefore, the differences and associations should be interpreted as subtle but potentially consequential at the population level. Third, because participation was voluntary, the sample may overrepresent individuals with a particular interest in diversity, equity, or bias, introducing potential self-selection bias. Finally, the fact that the physical disability IAT on the Implicit Project website uses identity-first language (e.g., “disabled people”) may be seen as a limitation because person-first language (i.e., “people with disabilities”) has traditionally been promoted as a way to reduce stigma.^31^ However, recent literature suggests that person-first language in scientific writing may actually increase rather than decrease stigma.^32^ Moreover, policies mandating the use of person-first language overlook the diverse language preferences among disabled people, including disabled researchers.^33^ Accordingly, the American Psychological Association (APA) now states that “both person-first and identity-first approaches to language are designed to respect disabled persons; both are fine choices overall”.^34^

### Conclusions

This study examines the evolution of implicit and explicit attitudes toward people with disabilities over time among male and female clinicians, rehabilitation assistants, and individuals in other professions. By analyzing this evolution over nearly two decades, this study provides novel insights into temporal patterns of implicit and explicit biases. The contrast between the evolution of implicit and explicit attitudes suggests that implicit biases remain resistant to change despite increased societal awareness and positive consideration of people with disabilities. Understanding these patterns may guide decision-making and help prioritize interventions and training programs aimed at reducing bias among healthcare practitioners.

## DECLARATIONS

### Ethical approval

Project Implicit was approved by the Institutional Review Board for the Social and Behavioral Sciences at the University of Virginia, USA, and the current study was approved by the University of Ottawa Research Ethics Board (H-02-25-11349), Canada.

### Data and Code Availability

In accordance with good research practices,^35^ the R scripts used to analyze the data are publicly available in Zenodo.^36^ The dataset and materials for the Implicit Association Test are available in the Project Implicit Demo Website Datasets, hosted on the Open Science Framework (OSF).^18^ This manuscript was published prior to peer review in the MedRxiv preprint repository on March 13, 2024.^37^

### Artificial Intelligence

ChatGPT (OpenAI) and DeepL were used to refine the language and improve readability of this manuscript.

### Reporting Guidelines

This manuscript conforms to the STROBE guidelines for observational studies.^38^

## Data Availability

In accordance with good research practices (Boisgontier, 2022), the R script used to analyze the data are publicly available in Zenodo (Boisgontier, 2025). The dataset and materials for the Implicit Association Test are available in the Project Implicit Demo Website Datasets, hosted on the Open Science Framework (OSF) (Xu et al., 2025).
References:
– Boisgontier MP. Research integrity requires to be aware of good and questionable research practices. Eur Rehabil J. 2022;2(1):1-3. https://doi.org/10.52057/erj.v2i1.24
– Boisgontier MP. Evolution of attitudes toward people with disabilities in healthcare practitioners and other occupations from 2006 to 2024: material, data, and R script [Data set]. Zenodo. 2025. https://doi.org/10.5281/zenodo.15008783
– Xu FK, Lofaro N, Nosek BA, Greenwald AG, Axt J, Simon L, Frost N, O'Shea B. Project Implicit Demo Website Datasets / Disability IAT 2004-2024 [dataset]. University of Virginia, Center for Open Science. Published August 7, 2014. Updated January 12, 2025. Accessed March, 2025. https://doi.org/10.17605/OSF.IO/Y9HIQ

## SUPPLEMENTARY MATERIAL

**Suppl. Material 1.** Procedures of the Implicit Association Test (IAT)

**Suppl. Material 2.** Explicit Attitudes

**Suppl. Material 3.** Occupation categories

**Suppl. Material 4.** Number, age, and sex of participants per year and by occupation group

**Suppl. Material 1.** Procedures of the Implicit Association Test (IAT) Participants completed a series of categorization tasks, totaling 180 trials, in which they sorted words and images into groups by pressing designated keys on a keyboard. The categories appeared on the left and right sides of the computer screen, and participants were instructed to press the “E” key if the presented stimulus belonged to the left-side category and the “I” key if it belonged to the right-side category. Participants were asked to respond as quickly and accurately as possible. If a participant placed a stimulus in the incorrect category, a red “X” appeared on the screen, and the correct response had to be selected before proceeding. In the General Disability IAT, participants performed seven sequential blocks (Fig. 1A): (1) Participants categorized the images (Fig. 1B) of people with or without disabilities into the respective categories: “disabled people” and “abled people”. (2) Participants categorized 16 words (Fig. 2C) into evaluative attribute categories (good vs. bad). (3) The disability and attribute categories were paired for 20 trials. For example, “disabled people” and “good” shared the same response key, while “abled people” and “bad” shared the other key. (4) The third block was repeated with 40 additional trials. (5) Similar to the first block of 20 trials but “disabled people” and “abled people” switched sides. (6) Similar to the third block of 20 trials but with a different pairing (e.g., “disabled people” and “bad” shared the same response key, while “abled people” and “good” shared the other key). (7) The sixth block was repeated with 40 additional trials. In the Physical Disability IAT, the word “physically” was added before each instance of the words “abled” and “disabled” (e.g., “physically disabled people”). Before each block, participants were provided detailed on-screen instructions, explaining the category pairing for the upcoming block and emphasizing the need for speed and accuracy. The same 6 images were used for each target concept across series (Fig. 1B-D). For each series, a set of 8 words was randomly selected from a set of 16 words for each attribute (Fig. 1C). Implicit attitudes toward people with and without disabilities were assessed using the D-score measure, which typically ranges from about –2 to 2. A positive D-score indicates that participants responded faster on stereotype-consistent trials than on stereotype-inconsistent trials, reflecting an implicit preference for people without disabilities. A negative D-score indicates the opposite, reflecting an implicit preference for people with disabilities. Absolute D-scores are interpreted as follows: no implicit preference (|D| < 0.15), slight implicit preference (0.15 ≤ |D| <0.35), moderate implicit preference (0.35 ≤ |D| < 0.65), and strong implicit preference (|D| ≥ 0.65).

**Suppl. Material 2.** Explicit Attitudes Explicit attitudes were assessed using a 7-point Likert scale in which participants rated their preference for people with or without disabilities. A score of 1 indicated a strong preference for people with disabilities, 4 indicated no preference, and 7 indicated a strong preference for people without disabilities. Specifically, in the General Disability IAT, this measure was based on the question “Which statement best describes you?”, with response options as follows: (1) “I strongly prefer disabled persons to abled persons”, (2) “I moderately prefer disabled persons to abled persons”, (3) “I slightly prefer disabled persons to abled persons”, (4) “I like disabled persons and abled persons equally”, (5) “I slightly prefer abled persons to disabled persons”, (6) “I moderately prefer abled persons to disabled persons”, and (7) “I strongly prefer abled persons to disabled persons” (Fig. 2D). In the Physical Disability IAT, the word “physically” was added before each instance of the word “abled” and “disabled” (e.g., “I strongly prefer physically disabled people to physically abled people”) and the word “persons” was replaced by “people”.

**Suppl. Material 3.** Occupation categories.

|  |  |  |
| --- | --- | --- |
| Administrative Support - Supervisors | Food Service - Supervisors | Protective services - Law Enforcement |
| Administrative Support - Financial Clerks | Food Service - Cooks and food prep | Protective Services - Other (e.g., security, lifeguards, crossing guards) |
| Administrative Support - Information and Records | Food Service - Servers | Repair/Installation - Supervisors |
| Administrative Support - Recording, Scheduling, Dispatching, Distributing | Food Service - Other food service workers (e.g., dishwasher, host) | Repair/Installation - Electrical and Electronic |
| Administrative Support - Secretaries and Assistants | **Healthcare - Diagnosing and Treating Practitioners (MD, Dentist, etc.)** | Repair/Installation - Vehicle and Mobile Equipment |
| Administrative Support - Other Support (data entry, office clerk, proofreaders) | Healthcare - Technologists and Technicians | Repair/Installation - Other |
| Arts/Design/Entertainment/Sports - Art and Design | Healthcare - Nursing and Home Health Assistants | Retired |
| Arts/Design/Entertainment/Sports - Entertainers and Performers | **Healthcare - Occupational and Physical Therapist Assistants** | Sales - Supervisors |
| Arts/Design/Entertainment/Sports - Media and communication | Healthcare - Other healthcare support | Sales - Retail |
| Arts/Design/Entertainment/Sports - Media Equipment workers | Homemaker or Parenting | Sales - Sales Representatives and Services |
| Business - Business Operations | Legal - Lawyers, Judges, and related workers | Sales - Wholesale and Manufacturing |
| Business - Financial Specialists | Legal - Legal support workers | Sales - Other sales (e.g., telemarketers, real estate) |
| Computer/Math - Computer Specialists | Maintenance - Building and Grounds Supervisors | Science - Life Scientists |
| Computer/Math - Math Scientists | Maintenance - Building workers | Science - Physical scientists |
| Computer/Math - Math Technicians | Maintenance - Grounds Maintenance | Science - Social Scientists |
| Construction/Extraction - Supervisors | Management - Top Executives | Science - Life, Physical, Social Science Technicians |
| Construction/Extraction - Construction Trades | Management - Advertising, Sales, PR, Marketing | Service and Personal Care - Supervisors |
| Construction/Extraction - Helpers, Construction Trades | Management - Operations Specialists | Service and Personal Care - Animal Care |
| Construction/Extraction - Extraction (e.g., mining, oil) | Management - Other Management Occupations | Service and Personal Care - Entertainment attendants |
| Construction/Extraction - Other | Military - Officer and Tactical Leaders/Managers | Service and Personal Care - Funeral Service |
| Education - Postsecondary Teachers | Military - First-line enlisted supervisor/manager | Service and Personal Care - Personal Appearance |
| Education - Primary, Secondary, and Special Ed Teachers | Military - enlisted tactical, air/weapons, crew, other | Service and Personal Care - Transportation, Tourism, Lodging |
| Education - Other teachers and instructors | Production - Supervisors | Service and Personal Care - Other service (e.g., child care, fitness) |
| Education - Librarians, Curators, Archivists | Production - Assemblers and Fabricators | Social Service - Counselors, Social Workers, Community specialists |
| Education - Other education, training, and library occupations | Production - Food processing | Social Service - Religious Workers |
| Education - Student | Production - Metal and Plastic | Transportation - Supervisors |
| Engineers/Architects - Architects, Surveyors, Cartographers | Production - Printers | Transportation - Air Transportation |
| Engineers/Architects - Engineers | Production - Textile, Apparel, Furnishings | Transportation - Motor Vehicle Operators |
| Engineers/Architects - Drafters, Engineering and Mapping Technicians | Production - Woodworkers | Transportation - Rail Transport |
| Farming, Fishing, Forestry - Supervisors | Production - Plant and System Operators | Transportation - Water Transport |
| Farming, Fishing, Forestry - Agriculture | Production - Other | Transportation - Material Moving |
| Farming, Fishing, Forestry - Fishing and Hunting | Protective Service - Supervisors | Transportation - Other |
| Farming, Fishing, Forestry - Forest, Conservation, Logging | Protective Services - Fire fighting and prevention | Unemployed |
| Farming, Fishing, Forestry - Other |  |  |

This variable, which is central to our study, was added to the Project Implicit website in 2006. Therefore, the data included in the models range from 2006 to 2024.

**Suppl. Material 4.**
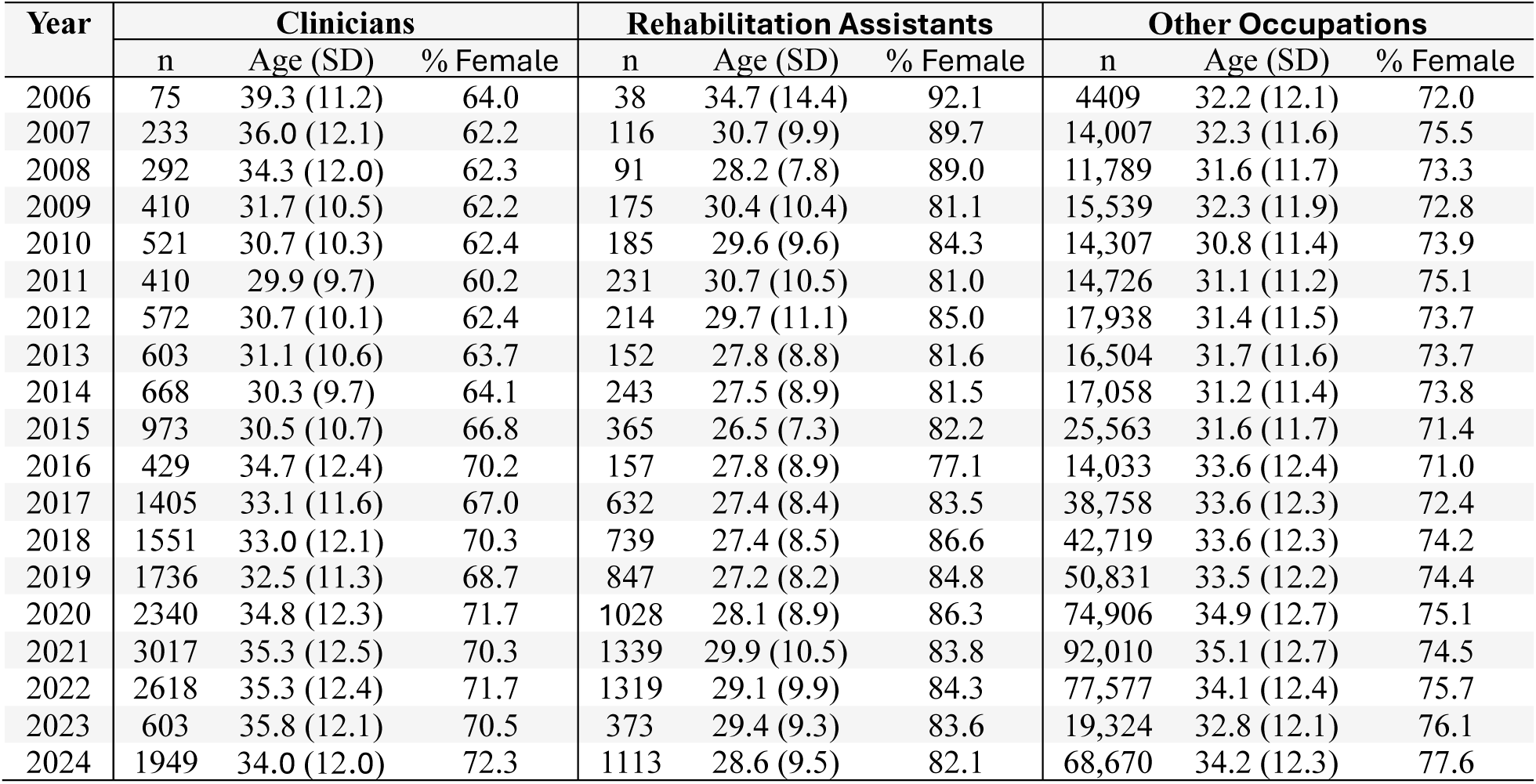
Number, age, and sex of participants per year and by occupation group.

| Year | Clinicians |  |  | Rehabilitation Assistants |  |  | Other Occupations |  |  |
| --- | --- | --- | --- | --- | --- | --- | --- | --- | --- |
|  | n | Age (SD) | % Female | n | Age (SD) | % Female | n | Age (SD) | % Female |
| 2006 | 75 | 39.3 (11.2) | 64.0 | 38 | 34.7 (14.4) | 92.1 | 4409 | 32.2 (12.1) | 72.0 |
| 2007 | 233 | 36.0 (12.1) | 62.2 | 116 | 30.7 (9.9) | 89.7 | 14,007 | 32.3 (11.6) | 75.5 |
| 2008 | 292 | 34.3 (12.0) | 62.3 | 91 | 28.2 (7.8) | 89.0 | 11,789 | 31.6 (11.7) | 73.3 |
| 2009 | 410 | 31.7 (10.5) | 62.2 | 175 | 30.4 (10.4) | 81.1 | 15,539 | 32.3 (11.9) | 72.8 |
| 2010 | 521 | 30.7 (10.3) | 62.4 | 185 | 29.6 (9.6) | 84.3 | 14,307 | 30.8 (11.4) | 73.9 |
| 2011 | 410 | 29.9 (9.7) | 60.2 | 231 | 30.7 (10.5) | 81.0 | 14,726 | 31.1 (11.2) | 75.1 |
| 2012 | 572 | 30.7 (10.1) | 62.4 | 214 | 29.7 (11.1) | 85.0 | 17,938 | 31.4 (11.5) | 73.7 |
| 2013 | 603 | 31.1 (10.6) | 63.7 | 152 | 27.8 (8.8) | 81.6 | 16,504 | 31.7 (11.6) | 73.7 |
| 2014 | 668 | 30.3 (9.7) | 64.1 | 243 | 27.5 (8.9) | 81.5 | 17,058 | 31.2 (11.4) | 73.8 |
| 2015 | 973 | 30.5 (10.7) | 66.8 | 365 | 26.5 (7.3) | 82.2 | 25,563 | 31.6 (11.7) | 71.4 |
| 2016 | 429 | 34.7 (12.4) | 70.2 | 157 | 27.8 (8.9) | 77.1 | 14,033 | 33.6 (12.4) | 71.0 |
| 2017 | 1405 | 33.1 (11.6) | 67.0 | 632 | 27.4 (8.4) | 83.5 | 38,758 | 33.6 (12.3) | 72.4 |
| 2018 | 1551 | 33.0 (12.1) | 70.3 | 739 | 27.4 (8.5) | 86.6 | 42,719 | 33.6 (12.3) | 74.2 |
| 2019 | 1736 | 32.5 (11.3) | 68.7 | 847 | 27.2 (8.2) | 84.8 | 50,831 | 33.5 (12.2) | 74.4 |
| 2020 | 2340 | 34.8 (12.3) | 71.7 | 1028 | 28.1 (8.9) | 86.3 | 74,906 | 34.9 (12.7) | 75.1 |
| 2021 | 3017 | 35.3 (12.5) | 70.3 | 1339 | 29.9 (10.5) | 83.8 | 92,010 | 35.1 (12.7) | 74.5 |
| 2022 | 2618 | 35.3 (12.4) | 71.7 | 1319 | 29.1 (9.9) | 84.3 | 77,577 | 34.1 (12.4) | 75.7 |
| 2023 | 603 | 35.8 (12.1) | 70.5 | 373 | 29.4 (9.3) | 83.6 | 19,324 | 32.8 (12.1) | 76.1 |
| 2024 | 1949 | 34.0 (12.0) | 72.3 | 1113 | 28.6 (9.5) | 82.1 | 68,670 | 34.2 (12.3) | 77.6 |

